# A CNN Autoencoder for Learning Latent Disc Geometry from Segmented Lumbar Spine MRI

**DOI:** 10.1101/2025.02.28.25323111

**Authors:** Mattia Perrone, D’Mar Moore, Daisuke Ukeba, John T. Martin

## Abstract

**Purpose:** Low back pain is the world’s leading cause of disability and pathology of the lumbar intervertebral discs is frequently considered a driver of pain. The geometric characteristics of intervertebral discs offer valuable insights into their mechanical behavior and pathological conditions. In this study, we present a convolutional neural network (CNN) autoencoder to extract latent features from segmented disc MRI. Additionally, we interpret these latent features and demonstrate their utility in identifying disc pathology, providing a complementary perspective to standard geometric measures.

**Methods:** We examined 195 sagittal T1-weighted MRI of the lumbar spine from a publicly available multi-institutional dataset. The proposed pipeline includes five main steps: 1) segmenting MRI, 2) training the CNN autoencoder and extracting latent geometric features, 3) measuring standard geometric features, 4) predicting disc narrowing with latent and/or standard geometric features and 5) determining the relationship between latent and standard geometric features.

**Results:** Our segmentation model achieved an IoU of 0.82 (95% CI: 0.80–0.84) and DSC of 0.90 (95% CI: 0.89–0.91). The minimum bottleneck size for which the CNN autoencoder converged was 4×1 after 350 epochs (IoU of 0.9984 - 95% CI: 0.9979–0.9989). Combining latent and geometric features improved predictions of disc narrowing compared to using either feature set alone. Latent geometric features encoded for disc shape and angular orientation.

**Conclusions:** This study presents a CNN-autoencoder to extract latent features from segmented lumbar disc MRI, enhancing disc narrowing prediction and feature interpretability. Future work will integrate disc voxel intensity to analyze composition.

## Introduction

Low back pain ranks among the most prevalent musculoskeletal conditions worldwide. In 2020, 619 million people were estimated to be impacted by low back pain and projections indicate that this number will surge to 843 million by 2050 [1]. Lumbar disc degeneration is associated with low back pain [2], however the factors that drive disc degeneration and pain in an individual patient are not well understood. Geometric characteristics of intervertebral discs are recognized in the progression of disc degeneration. For example, disc narrowing is a hallmark feature of degeneration identified on clinical imaging [3]. Furthermore, discs with a smaller ratio of disc area to disc height are prone to degeneration [4]. and local variations in disc geometry affect stresses at the disc-to-bone interface, which may lead to disc degeneration [5]. Thus, a thorough analysis of disc geometry may reveal underlying drivers of degeneration and low back pain.

Machine learning is often used to identify disc pathologies from imaging data [6][7][8][9]. Most algorithms implement convolutional neural networks (CNN) to perform radiological feature classification [10] including disc degeneration [11][12]. Such models achieve accuracy comparable to that of expert radiologists [10][13], however the features learned by the model often lack interpretability or are not evaluated and consequently are not explainable to clinicians. Clinicians are more likely to trust and adopt AI tools when they comprehend their decision-making process [14], as understanding how a model reaches its conclusions supports informed decision-making [15]. Modeling approaches leveraging tabular patient demographics and extracted disc geometry have also been developed [16][17][18]. While this ensures interpretability of the predictive features, these models have lower performance than models trained on image data, as the latter exploit a greater amount of information during training [6]. Thus, developing a deep learning model with explainable features may improve the adoption of a model evaluating spine health in clinical practice.

Autoencoders are among the most common models used for computer vision tasks, from denoising [19][20] to generating new samples [21][22]. As the autoencoder architecture allows for mapping input data into a lower dimensional space, feature learning is a main application for autoencoders. For example, autoencoders are reliable models for dimensionality reduction and outperform linear [23] and nonlinear [24][25] dimensionality reduction techniques. In spine research, autoencoders have been applied for automatic diagnosis of degenerative disc diseases. Specifically, extracting disc features in MRI using an autoencoder outperformed other manually-extracted geometric features when predicting disc pathology [26]. Similarly, sparse autoencoders and diffusion autoencoders have been employed to enhance interpretability and identify critical features in spinal imaging [27][28]. While these studies have demonstrated the effectiveness of autoencoders for feature extraction and prediction in spinal imaging, no studies have explored the relationship between these extracted latent features and geometric properties of the disc or the utility in using latent features to identify degenerative disc phenotypes.

This study introduces a CNN-autoencoder to extract features from segmented lumbar disc MRI for use in predicting disc narrowing, a common spinal pathology relevant to low back pain [29][30][31]. We applied our model directly to image masks to isolate the role that spinal geometry plays in driving pathology. We hypothesized that integrating a latent geometric representation with standard geometric determinants (i.e., disc height, anteroposterior width, lateral width and disc inclination angles) will enhance disc narrowing prediction beyond the use of standard geometric measures alone. Furthermore, we provided insight into the latent features by evaluating their relationship to disc geometry.

## Methods

### 2.1 Overview

We developed a computational pipeline **(Fig. 1)** to automatically segment intervertebral discs in lumbar spine MRI and to extract latent geometric features from each segmented disc. The computational pipeline included five steps: (1) MRI volumes were segmented using a Swin Transformer to generate disc masks; (2) disc masks served as inputs to a CNN-autoencoder for unsupervised feature extraction; (3) standard geometric features were calculated from disc masks (4); a gradient boosting classifier was trained on latent and standard disc geometric features to predict disc narrowing; (5) a statistical evaluation was performed to determine the relationship between latent and standard geometric features.

**Fig. 1.**
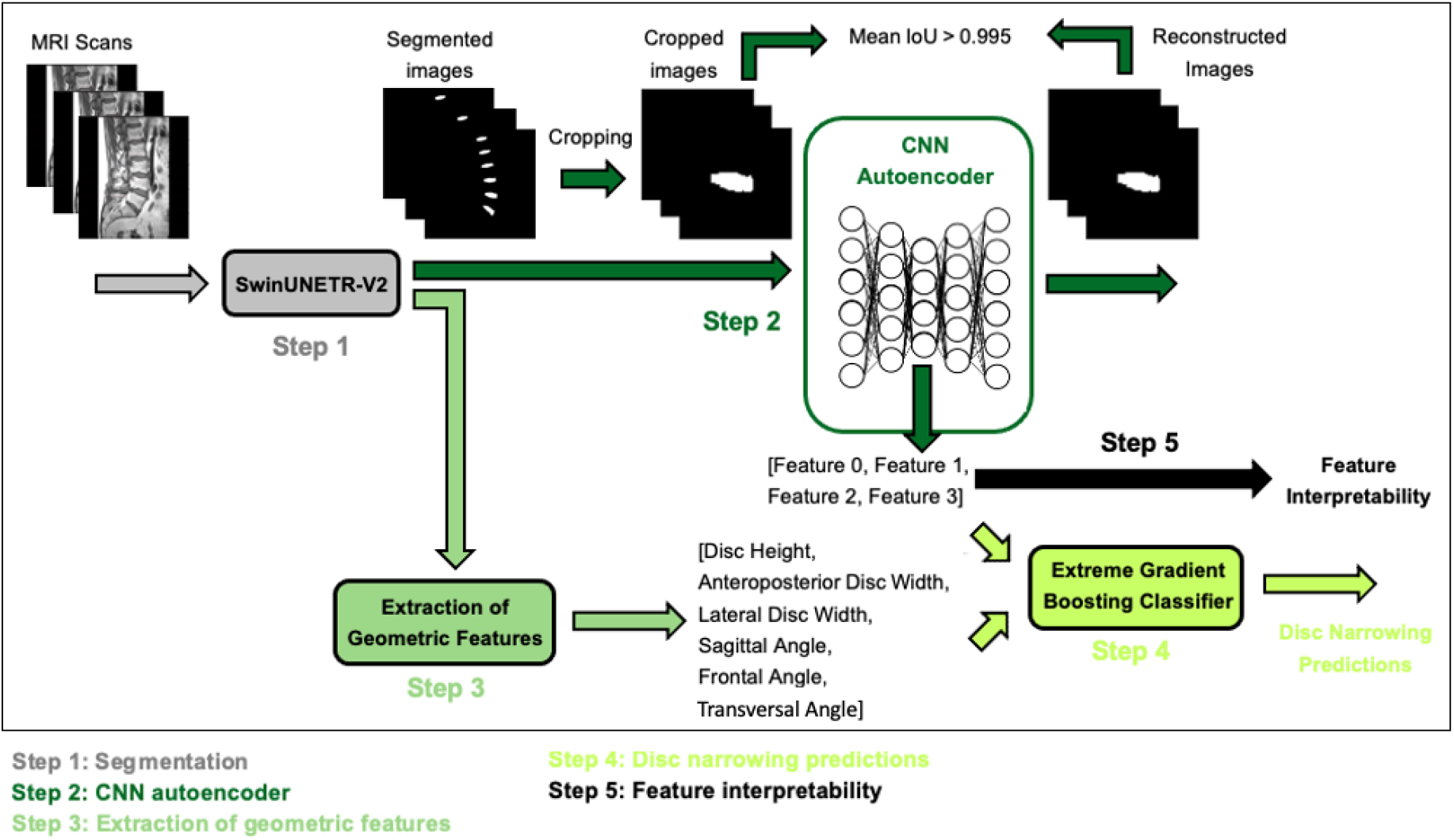
Pipeline followed in the current study

We leveraged a public dataset comprised of 195 sagittal T1-weighted lumbar spine MRI scans from 218 patients with chronic low back pain (N=75 male, 120 female) [32]. Imaging data were collected from different institutions (make: Siemens, Philips; field strength: 1.5-3T; TE: 8ms-124ms; TR: 446-5570ms; flip angle: 80-160 deg), resulting in a heterogeneous dataset to generalize across diverse imaging settings. For each patient, pixel-wise annotation of lumbar vertebrae, intervertebral discs and spinal cord is included. Each image was additionally annotated by a practicing spine surgeon (DU) to label disc narrowing.

### 2.2 Segmentation model

For MRI segmentation we used SwinUNETR-V2, a shifting window transformer model with convolutional blocks that enhance local feature extraction, achieving state-of-the-art performance on multiple 3D biomedical imaging datasets [33]. We divided the dataset allocating 80% (n=156) for training and validation and 20% for testing (n=39). Preprocessing of MRI scans included normalization of pixel values and image resampling to a resolution of 1.4 x 1.2 x 1.5 mm. The model was trained for 200 epochs with batch size=4, incorporating an early stopping mechanism when the validation intersection over union (IoU) did not improve over 15 consecutive epochs. A custom loss function that includes terms for binary cross-entropy and dice similarity coefficient (DSC) was implemented for training. The two terms had equal weights in determining the final loss. The model with the highest validation IoU across all training epochs was retained for subsequent analysis.

### 2.3 CNN Autoencoder

For unsupervised feature extraction, a CNN-autoencoder was trained using test masks from the segmentation process. The CNN-autoencoder architecture comprises 3D convolutional and deconvolutional layers within the encoder and decoder and batch normalization layers to promote training stability (**Fig. 2**). Individual discs were isolated from each spine resulting in a training set with 202 disc volumes from 39 spines. All data were used to train the model. Each disc image was padded and/or cropped to standardize the dimensions to 64x64x64, ensuring the disc is centered in each sample. Binary cross-entropy was used as the loss function. Model convergence was defined when the IoU between input and output masks exceeded 0.995, indicating that the input mask reconstruction was nearly perfect. The autoencoder bottleneck layer dimension represents a crucial hyperparameter that defines the number of features extracted from the segmented image. The bottleneck layer size was tested with different dimensions (64×1, 8×1, 4×1 and 3×1) and the minimum effective bottleneck size (n=4) was selected to reduce multicollinearity among features and enhance their interpretability. The model converged after 350 epochs with batch size=4. No skip connections were included to guarantee that all the information of the segmented image was mapped into the latent space.

**Fig. 2.**
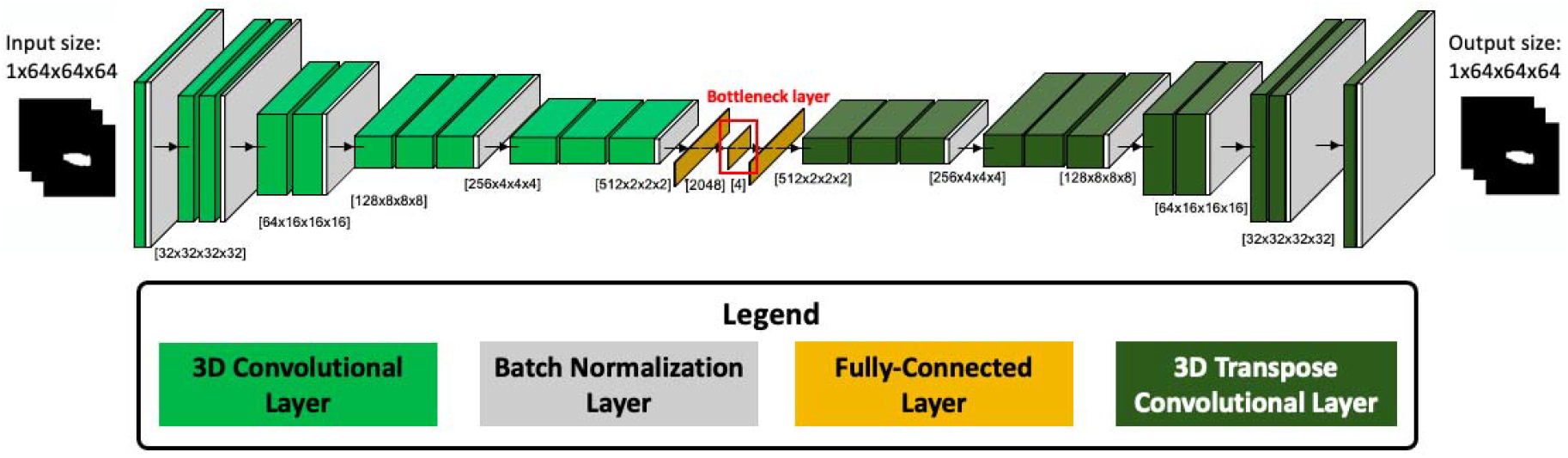
Autoencoder architecture used for unsupervised feature extraction

### 2.4 Extraction of geometric features

Geometric features were automatically extracted from segmented images including disc height, anteroposterior disc width and lateral disc width, as well as the inclination of the disc relative to the horizontal axis in the sagittal plane and to the vertical axis in the transverse and coronal plane (**Fig. 3**). Disc height was calculated as the mean height across all pixels within the segmented disc volume (sagittal plane), anteroposterior disc width as the distance between the two lateral edges of the segmented disc on the middle slice (sagittal plane) and lateral disc width as the distance between the two lateral edges of the segmented disc on the middle slice (transverse plane).

**Fig. 3.**
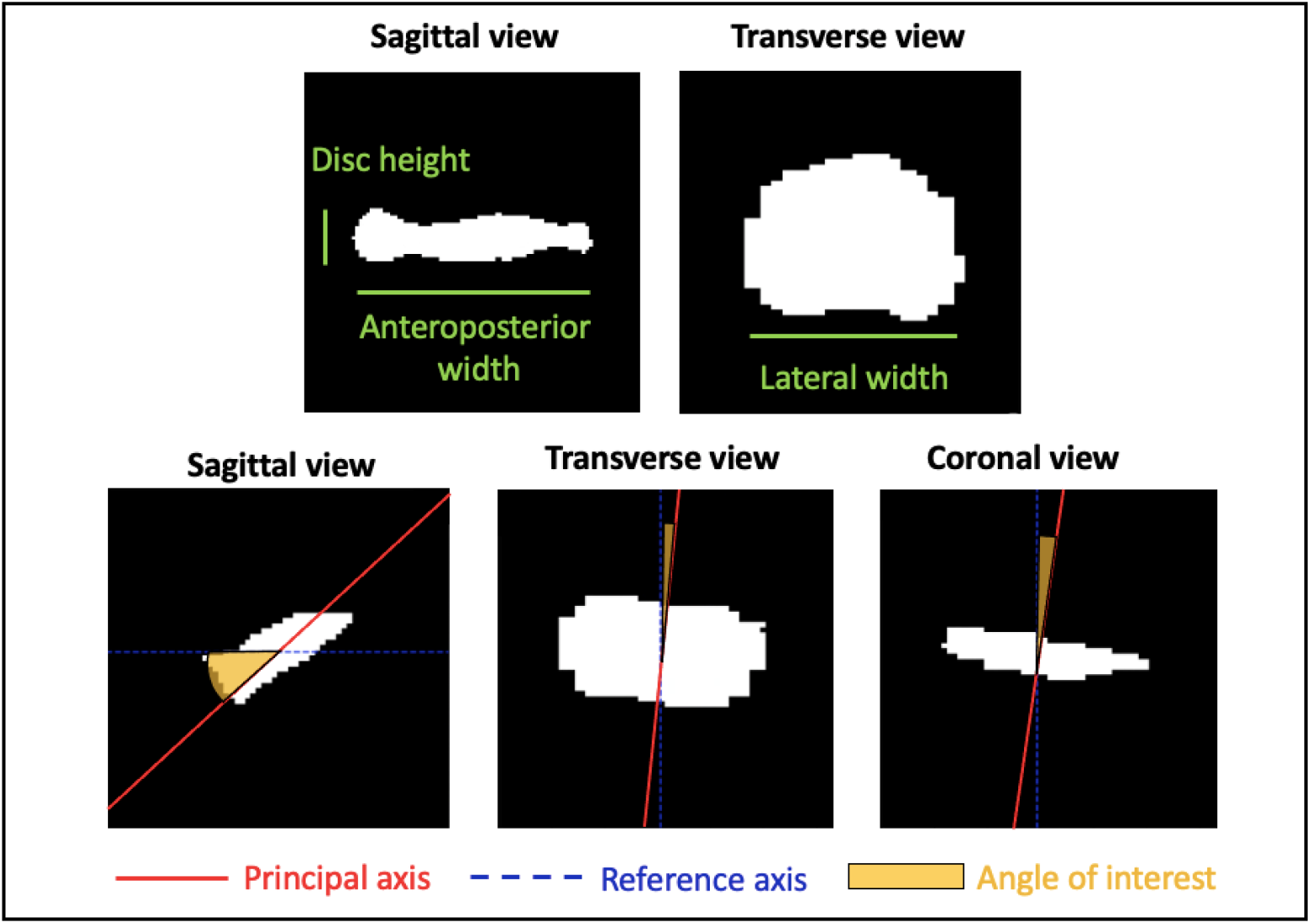
Visualization of geometric features extracted from lumbar disc masks: disc height, anteroposterior width and lateral width (top row), along with the angle of interest computed between the principal axis (red) and the reference axis (blue) in sagittal, transverse and coronal views (bottom row). All measurements except disc height were performed on the middle slice

A custom Python script was implemented to compute the angular measurements. This consists of identifying the principal axis of segmented discs using principal component analysis (PCA) on the pixel coordinates and calculating the angle of inclination relative to the horizontal or vertical axis, depending on the anatomical plane.

### 2.5 Gradient boosting

A gradient boosting classifier (XGBoost [34]) was implemented to predict disc narrowing (max estimator depth=3, number of estimators=200, learning rate=0.1). Since disc narrowing prediction is a binary classification problem, F1-score and area under the receiver operating characteristic curve (AUC ROC) were used as evaluation metrics. F1-score was specifically chosen to account for the data imbalance, as narrowed discs constitute approximately 20% of the dataset. 5-fold cross-validation was performed to ensure the reliability of the results obtained.

### 2.6 Feature interpretability

To interpret the latent features, we designed an experiment to systematically interrogate how latent features were related to 3D disc geometry. To do so, a synthetic dataset was generated by fixing three of the four latent features at their respective mean and varying the fourth feature from its minimum to maximum value. For each latent feature, predicted disc masks were generated for geometry measurements and visualization at the minimum, 1/5, 2/5, 3/5 and 4/5, and maximum of the range (**Fig. 4**).

**Fig. 4.**
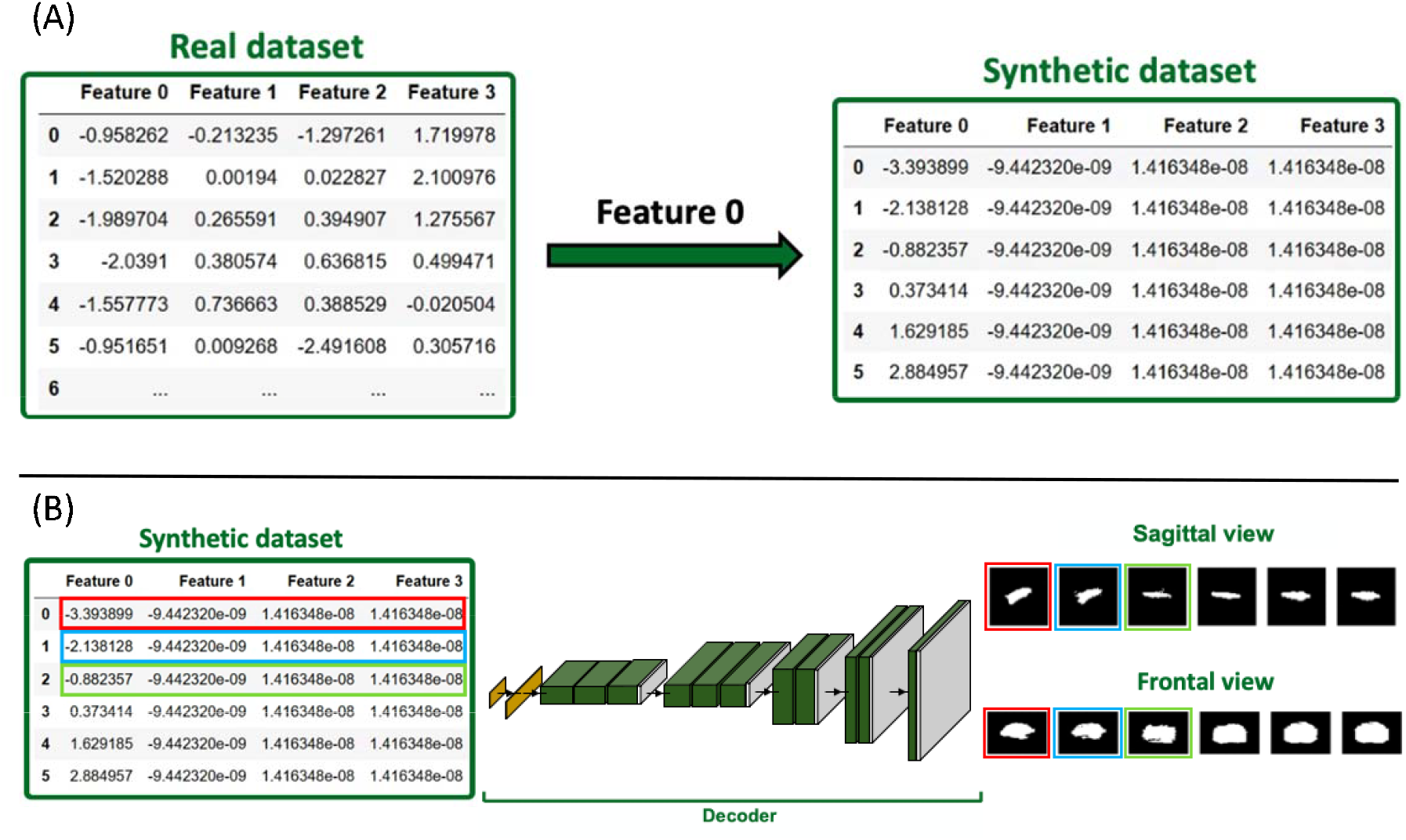
Overview of the interpretability analysis for latent features: (A) Generation of the synthetic latent feature dataset starting from the real latent feature dataset by varying latent features, with the process illustrated specifically for Feature 0. (B) Decoder architecture used to reconstruct images from the synthetic dataset, with examples of sagittal and frontal views of reconstructed masks. Input features and reconstructed images are marked with matching frame colors, indicating that the former serves as input to the decoder to produce the latter

## Results

For segmentation via the Swin Transformer, we obtained an Intersection over Union (IoU) of 0.82 (95% CI, 0.80-0.84) and Dice similarity coefficient (DSC) of 0.90 (95% CI, 0.89-0.91), which aligns to what reported in the literature for the same dataset [32]. The autoencoder achieved an IoU of 0.9981 (95% CI: 0.9976–0.9986) using a bottleneck size of 64×1 in 190 epochs. Reducing the bottleneck to 8×1 improved the IoU to 0.9983 (95% CI: 0.9979–0.9987) after 199 epochs, while further reduction to 4×1 resulted in the highest IoU of 0.9984 (95% CI: 0.9979–0.9989) in 350 epochs. In contrast, a bottleneck of 3×1 did not converge after 1000 epochs, with the IoU oscillating between 0.6 and 0.8. Based on these results, we selected a bottleneck size of 4×1 to minimize multicollinearity.

Results for predicting disc narrowing are reported in **Table 1** (ground truth masks) and **Table 2** (predicted masks). Disc narrowing predictions improved when combining geometric and latent features compared to using either feature set individually.

**Table 1.**
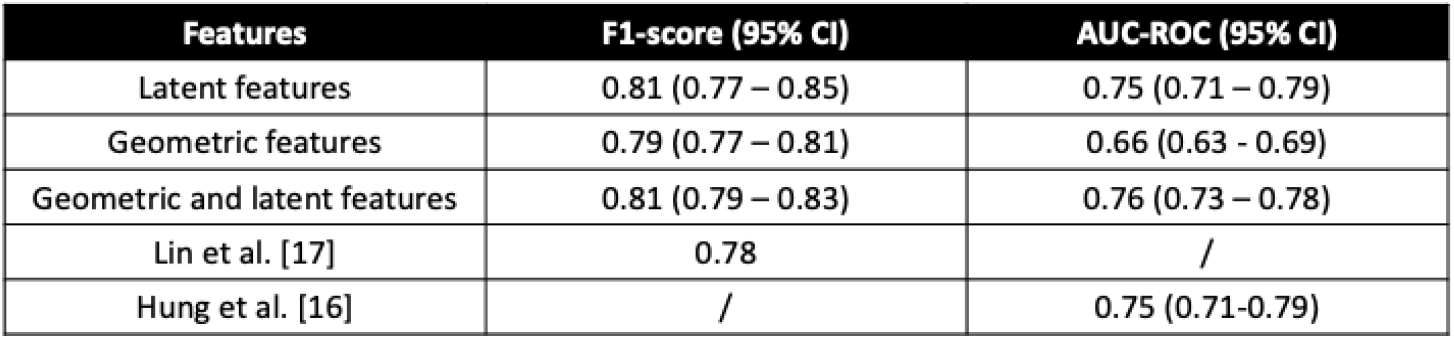
Predictions of disc narrowing using latent and geometric features from ground truth masks. Comparisons made to relevant examples in literature

| Features | F1-score (95% CI) | AUC-ROC (95% CI) |
| --- | --- | --- |
| Latent features | 0.81 (0.77 – 0.85) | 0.75 (0.71 – 0.79) |
| Geometric features | 0.79 (0.77 – 0.81) | 0.66 (0.63 – 0.69) |
| Geometric and latent features | 0.81 (0.79 – 0.83) | 0.76 (0.73 – 0.78) |
| Lin et al. [17] | 0.78 | / |
| Hung et al. [16] | / | 0.75 (0.71-0.79) |

**Table 2.** Predictions of disc narrowing using latent and geometric features from predicted masks.

| Features | F1-score (95% CI) | AUC-ROC (95% CI) |
| --- | --- | --- |
| Latent features | 0.77 (0.75 – 0.78) | 0.58 (0.54 – 0.62) |
| Geometric features | 0.79 (0.77 – 0.81) | 0.66 (0.63 – 0.69) |
| Geometric and latent features | 0.79 (0.77 – 0.81) | 0.69 (0.63 – 0.73) |

Finally, we analyzed the correlation between geometric and latent features using correlation coefficients to identify significant relationships between the two (**Fig. 5**). To do so, we generated a synthetic dataset and visualized how each latent feature influenced disc geometry (**Fig. 4** and **Fig. 5)**.

**Fig. 5.**
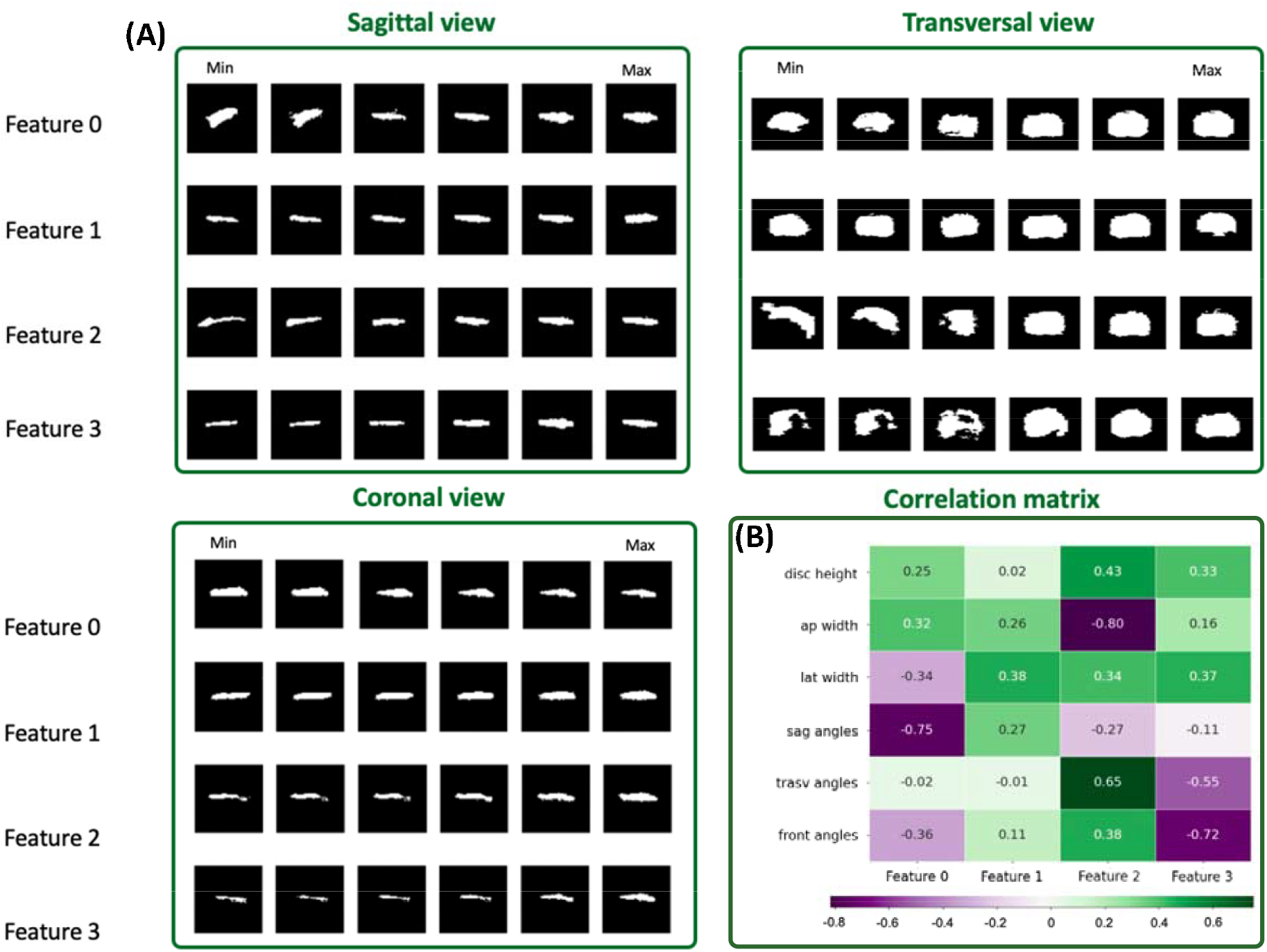
(A) Visualization of reconstructed masks across sagittal and axial views for different latent features, ranging from minimum to maximum values. For clarity, only the central slice in each plane is displayed. (B) Correlation analysis of latent features and geometric features

We then examined the relationship between latent and geometric features by calculating a correlation matrix and found that some latent features are strongly correlated with geometric features – e.g., Feature 2 and anteroposterior width (corr coeff=-0.80; p-value<0.001) and Feature 0 and sagittal angles (corr coeff=-0.75; p-value<0.001) (**Fig. 5**). Of note, latent features were not correlated with other latent features (corr coeff <0.05).

## Discussions

Disc geometry plays a key role in disc health and mechanical function [35][36]. To better understand disc geometry, we developed a CNN-autoencoder to extract latent features from segmented lumbar disc MRI and determine what geometry latent features encode and whether they were related to disc degeneration. We identified a minimum latent feature set size of 4 was capable of encoding disc geometry and that these latent features improved the prediction of disc narrowing over the use of geometric features alone (**Table 1** and **Table 2**). Furthermore, latent features encoded disc geometry including disc shape and angular orientation.

We identified strong correlations between latent features and disc shape (e.g., Feature 2 and anteroposterior width) and rotation angles (e.g., Feature 0 and sagittal angle), while correlations to other geometric features is weaker (disc height and lateral width). The autoencoder generates a set of orthogonal features that spans the latent space, which is informative for differentiating between discs. Additionally, only specific combinations of features reconstruct disc masks, as some images in **Fig. 5** fail to preserve the anatomical proportions of the discs (e.g., transversal view - row 3, column 1; transversal view - row 4, column 3). We surmise that some unnatural disc configurations exist in the latent feature space. When predicting disc narrowing using features extracted from deep learning-segmented images (**Table 2**), performance is inferior compared to using features derived from ground truth masks (**Table 1**), suggesting that improving segmentation accuracy may enhance overall prediction performance.

The first step of our computational pipeline is segmentation, a foundational task in medical imaging studies. We utilized a Swin Transformer model [37], which produces results (IoU: 0.82; DSC: 0.90) consistent with those reported by other groups using the same dataset (32). Importantly, the dataset used in this study comprises data from four different institutions, introducing heterogeneity that may reduce the segmentation model’s performance compared to studies using data from a single institution [38]. Our approach to dimensionality reduction for intervertebral disc geometry using a CNN-Autoencoder is similar to that of Peloquin et al. [39]. While these two studies use different methods to reduce segmented disc images to latent geometric features (deep learning versus statistical shape modeling; multiple spinal levels versus single level), both identify correlations between features and standard disc geometry measures. We surmise a parsimonious set of features can parameterize a wide range of disc geometry, revealing a surprising simplicity in disc shape.

Our approach to predict disc pathology using standard and latent geometric features produced similar results to models that do not use latent geometric features. Lin et al. [17] developed a classification model from disc height and anthropometrics (age, sex and BMI). Hung et al. [16] conducted a similar study, implementing two different models for the same prediction task: one leveraging anthropometrics (age, sex, body height and body weight) and another using both anthropometrics and geometric features (disc height and disc depth). Our model predictions incorporating latent and standard geometric features matched those of Lin et al. [17] and Hung et al. [16]. In both investigations, patient anthropometrics proved relevant for making predictions of disc pathologies. Anthropometric variables may further improve the performance of models including latent and standard geometric features. However, these were absent from the public dataset used in the current study.

This study introduces a CNN-autoencoder for extracting latent features from segmented MRI images of lumbar discs. These latent features provide complementary information to standard disc geometric features, as demonstrated by improved metrics in predicting disc narrowing. Furthermore, the proposed approach enhances feature interpretability by elucidating the relationship between latent and geometric features. Limitations of this study include the use of a dataset that is skewed towards cases of disc narrowing, potentially affecting the generalizability of the findings, which we mitigated by using F1-score to account for class skewness. Additionally, the segmentation model was limited to a single architecture, with no comparison to alternative approaches. Future work will explore integrating latent variables related to disc voxel intensity to incorporate valuable insights from disc composition and look at the relationship between latent features and clinical outcomes.

## Data Availability

All data produced in the present study are available upon reasonable request to the authors

## Statements and Declarations

This work was supported by the National Institutes of Health (NIH) under Grant R00AR077685. The authors have no conflicts of interest to declare

## Notes

### Competing Interest Statement

The authors have declared no competing interest.

### Funding Statement

This study did not receive any funding

### Author Declarations

The study used only openly available human data that were originally located at https://zenodo.org/records/8009680

